# The interplay between vaccination and social distancing strategies affects COVID19 population-level outcomes

**DOI:** 10.1101/2020.12.22.20248622

**Authors:** Sharon Guerstein, Victoria Romeo-Aznar, Ma’ayan Dekel, Oren Miron, Nadav Davidovitch, Rami Puzis, Shai Pilosof

## Abstract

Social distancing is an effective population-level mitigation strategy to prevent COVID19 propagation but it does not reduce the number of susceptible individuals and bears severe social consequences—a dire situation that can be overcome with the recently developed vaccines. Although a combination of these interventions should provide greater benefits than their isolated deployment, a mechanistic understanding of the interplay between them is missing. To tackle this challenge we developed an age-structured deterministic model in which vaccines are deployed during the pandemic to individuals who, in the eye of public health, are susceptible (do not show symptoms). The model allows for flexible and dynamic prioritization strategies with shifts between target groups. We find a strong interaction between social distancing and vaccination in their effect on the proportion of hospitalizations. In particular, prioritizing vaccines to elderly (60+) before adults (20-59) is more effective when social distancing is applied to adults or uniformly. In addition, the temporal reproductive number *R*_*t*_ is only affected by vaccines when deployed at sufficiently high rates and in tandem with social distancing. Finally, the same reduction in hospitalization can be achieved via different combination of strategies, giving decision makers flexibility in choosing public health policies. Our study provides insights into the factors that affect vaccination success and provides methodology to test different intervention strategies in a way that will align with ethical guidelines.

**Author summary:** A major question in epidemiology is how to combine intervention methods in an optimal way. With the recent deployment of COVID19 vaccine, this question is now particularly relevant. Using a data-driven model in which vaccines are deployed during the pandemic and their prioritization can shift between target groups we show that there is a strong interplay between these interventions. For example, prioritizing vaccines to elderly—the common strategy worldwide—results in a larger reduction in hospitalizations when social distancing is applied to adults than to elderly. Importantly, reduction in hospitalizations can be achieved via multiple combination of intervention strategies, allowing for flexible public health policies.

## Introduction

Vaccines are an essential tool for reducing the burden of endemic diseases, such as measles, hepatitis, and influenza. For such diseases, vaccine development and production is, by now, a well-practiced process. Nevertheless, developing, producing, and deploying vaccines in the midst of a pandemic is a great challenge. For example, during the H1N1 pandemic vaccines were limited and their deployment required prioritization [1]. COVID19 is a typical example of such a situation. Without vaccines, the only strategy to contain COVID19 is to cut transmission chains by reducing contacts. Public health measures such as social distancing, improved hygiene, and face masks effectively reduce the risk of infection but do not reduce susceptibility among the population. Moreover, social-distancing interventions also bear social, economic, and psychological consequences [2, 3]. Therefore, vaccine development for COVID19 has been carried out at an unprecedented pace. Nevertheless, even now, when a vaccine is available, its deployment requires prioritization due to high demand and low supply. Thinking ahead about relevant strategies for deployment may save not only valuable time for policy makers [1, 4], but eventually lives and it is therefore at the forefront of debate and research [5–9].

Silent infections, caused by presymptomatic and asymptomatic individuals, are a major driving force of COVID19. This is because interventions such as quarantine are applied when an individual shows symptoms. Moreover, vaccination is first prioritized to those individuals who have not been tested positive (identification of which depends on symptoms). The importance of silent infections is also reflected in modeling exercises. For example, Moghadas et al. [10] showed that attack rate decreases with the proportion of silent infections that are identified. In their model, attack rate can be brought to below 1% when about 40% of silent infections are identified.

Social distancing is the main strategy we currently have to deal with silent infections. Therefore, it is crucial to consider the interplay between vaccination and social distancing because, even when a vaccine is deployed, social-distancing is still enforced at some level. Several studies have examined vaccination strategies [6, 11–15], addressing multiple aspects of this issue. While these studies vary in their modeling frameworks, a general emerging result is that vaccines should be prioritized to the elderly and essential workers. Yet no study that we know of has explicitly tested the interplay between these two interventions in a realistic scenario in which vaccines are deployed according to a prioritization strategy. A mechanistic understanding of this interplay can serve to hone adequate strategies and understand possible caveats with a particular choice of strategy. Beyond COVID19, it addresses a general and fundamental question in epidemiology on how to mitigate disease with a combination of intervention strategies (e.g., antimalarial drugs and bed nets for vector control; [16]).

Here, we examine combinations of vaccine deployment and social distancing, and the interaction between them, using a data-driven, age-structured, deterministic model. We implement vaccination dynamics that reflect real-world scenarios by deploying vaccines to individuals who, in the eye of public health, are susceptible: presymptomatic and asymptomatic (exposed and infectious), those that have recovered from an asymptomatic infection, and susceptible individuals (those who were never infected). We model a scenario in which vaccines are deployed while social distancing is still in place—a highly likely situation in every country. Our goal is to illuminate qualitative effects of the interplay between vaccine deployment and social distancing. Which strategy combination can minimize adverse outcomes, and how? We focus on reduction in hospitalizations because hospital care is a limited resource in public health, which is also correlated with mortality. We show that the interplay between these interventions can have a synergistic effect. Moreover, the same reduction in hospitalization can be achieved via different combination of strategies, giving decision makers flexibility in choosing public health policies. These results were consistent for four countries.

## Results

We developed an age-structured deterministic compartmental model that reflects the main states of COVID19 progression, including a presymptomatic stage, which has been shown to affect transmission and possible control efforts [10, 17] (Fig. 1, Fig. S1). We used 10-yr interval age-grouping (0-9, 10-19,…,70-79,80+), in line with previous studies of COVID19 (e.g, [18]) and with the data management protocols of most countries. The model and choice of parameter values are detailed in the Methods.

**Fig 1.**
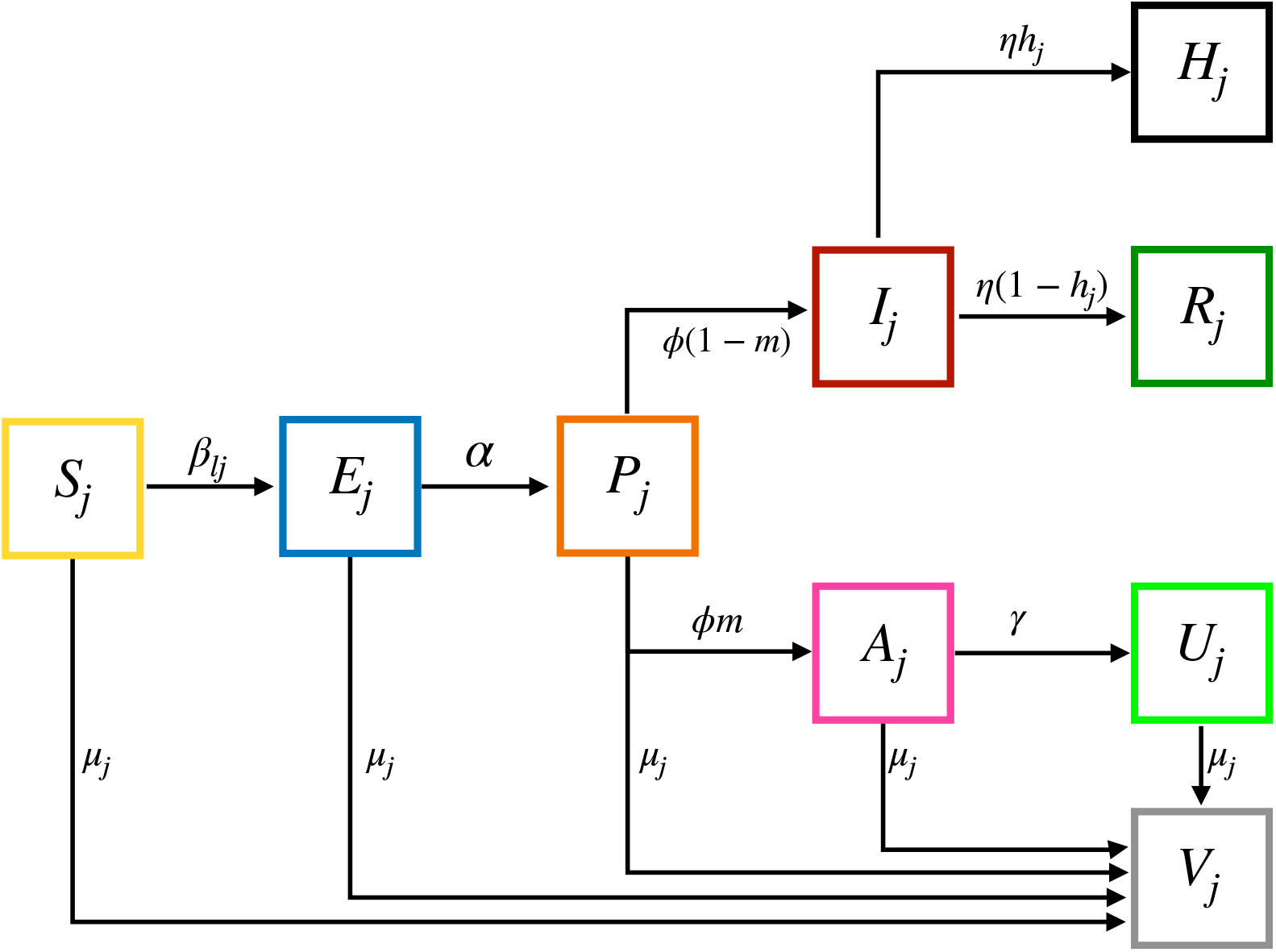
Model description. Individuals transition between states with rates specified by Greek letters. Small Latin letters are probabilities. Subscripts *j* and *l* depict age groups. Individuals start at a susceptible state (*S*) and upon infection with an age-dependent rate *β*_*lj*_ become infected with the virus (*E*). After an incubation period of *α* days individuals become presymptomatic (*P*) for *f* days and infectious. The disease can then progress to be either asymptomatic (*A*) with a probability *m*, or symptomatic (*I*). Symptomatic individuals are identified and removed to quarantine (*R*) within *η* days or, with a given age-dependent probability *h*_*j*_ develop severe symptoms and go to the hospital (*H*). Asymptomatic individuals naturally recover (*U*) within *γ* days. All individuals that do not show symptoms can get a vaccine (*V*) at a constant rate of *µ* vaccines per day. See Methods for a comprehensive description of the model equations, parameters and vaccination strategies. See examples of model run in (Figs. S1,S2).

Vaccines are deployed at a constant rate of *κ* vaccines per day, which we measure as the percentage of population that a government can vaccinate a day (e.g., *κ* = 0.2 translates to deployment of 16,000 vaccines per day in a population of 8 million people). The amount of daily vaccines is divided proportionally to the size of the target age groups. Following [19] we apply an 80% vaccination acceptance proportion for the target groups. We explore two vaccination strategies: (i) ‘elderly first’, in which elderly (ages 60+) are vaccinated first and adults (20-59) next, and (ii) ‘adults first’, in which adults are vaccinated before elderly (Fig. S3). The elderly first strategy is currently being applied in most countries that have started vaccination campaigns (e.g., Israel). Once the prioritized age group is fully vaccinated, the vaccines are applied to the next group (Figs. S2,S3). Vaccines are currently not developed for children and we therefore did not include ages 0-19 in vaccination strategies.

We present results in the main text for Israel, a small country of ≈8.7 million people, but we also tested our model using age demographics and contact matrices for Italy, Belgium and Germany, and results were qualitatively the same for all countries (see Sensitivity Analyses in Methods).

### Social distancing affects vaccination strategies non-linearly

A combination of social distancing and vaccination is expected to have a synergistic, stronger effect in decreasing hospitalizations than any of these measures alone, and this effect should increase with stronger social distancing and higher vaccination rates. However, it is unclear how this interplay will be affected by vaccination rates and the strategy of social distancing. To test this, we apply social distancing either uniformly, to all the population regardless of age, or in a targeted way towards elderly or adults. We impose social distancing by reducing contact rates of specific ages (see Methods). We measure the impact of vaccine deployment as the percent of reduction in hospitalizations (in all the population) compared to a no-vaccination scenario as

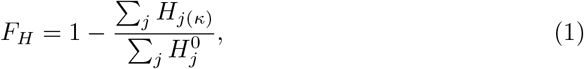

where *H*_*j*(*κ*)_ is the total cumulative number of hospitalizations in age group *j* for a given deployment rate *κ*, and 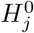 is the total number of hospitalizations in age group *j* when a vaccine is not deployed (Fig. S4).

As expected, vaccine impact increases with vaccination rates (line colors in Fig. 2). Prioritizing vaccines to adults does not reduce hospitalizations as effectively as when prioritizing to elderly (compare left to right columns in Fig. 2). However, there is a strong interaction between vaccination and social distancing strategies in their effects on *F*_*H*_. Combining an ‘elderly first’ strategy with adult-targeted or uniform social distancing increases *F*_*H*_ in a non-linear, synergistic way (Fig. 2A,C), while targeting both interventions at the elderly is not as effective (Fig. 2B). In fact, when both interventions are targeted at the same group (elderly or adults), *F*_*H*_ varies very little or decreases with increasing strength of social distancing (Fig. 2B,D). This is a result of an overly-effective social distancing, which by itself prevents hospitalizations, overriding the need for vaccinations. This phenomenon also explains the decrease in *F*_*H*_ when social distancing is applied to all the population too strongly (Fig. 2C,F).

**Fig 2.**
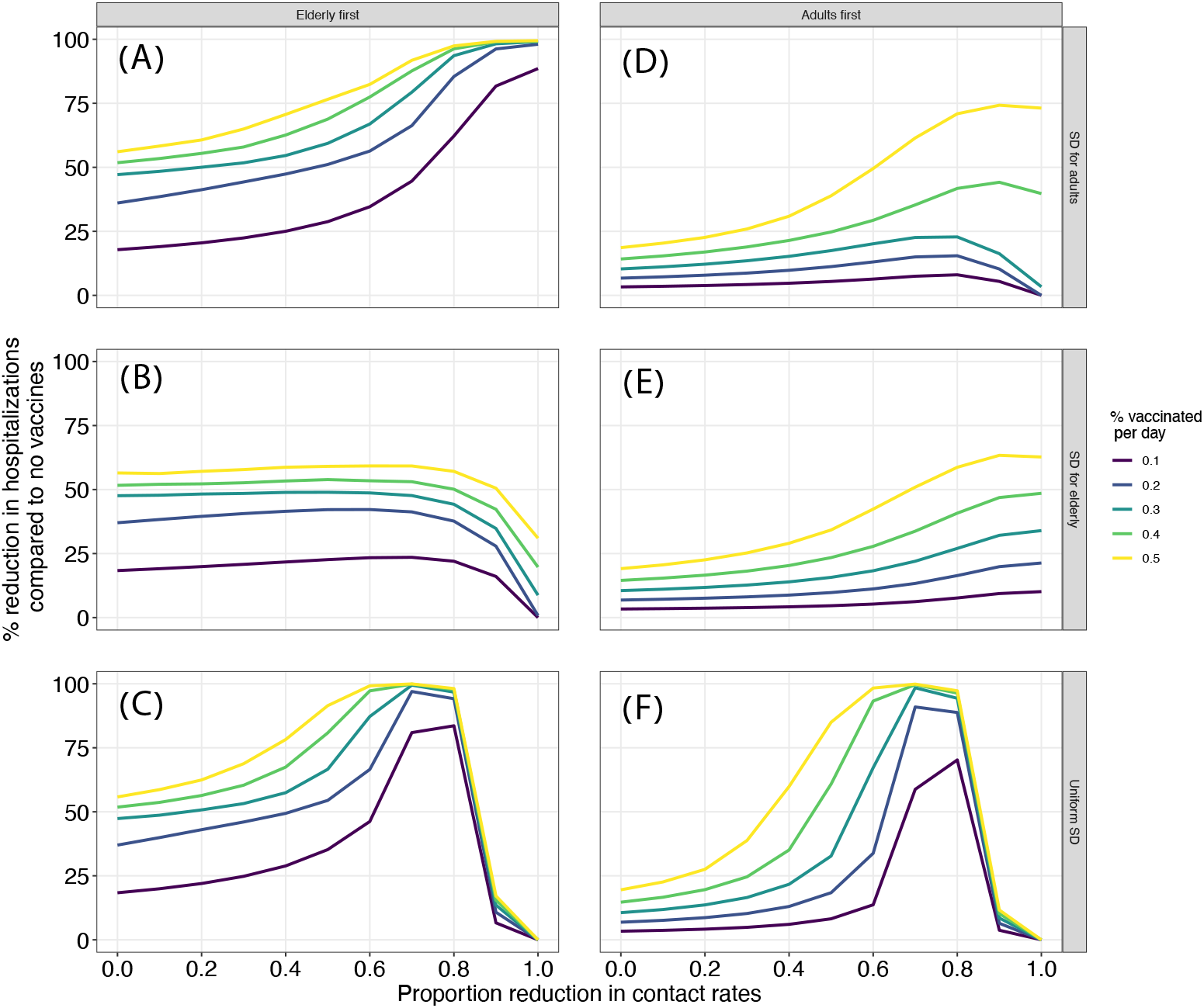
Effect of joint interventions on vaccination efficiency. The plot depicts *F*_*H*_ (y-axis) as a function of strength of social distancing (x-axis), vaccination rates (*κ*; line colors), vaccination strategies (columns) and social distancing strategies (rows).

### Proportion hospitalized is determined by the interplay between intervention strategies

Because we find that social distancing can affect vaccination deployment efficiency, we now turn to quantify the joint effects of these interventions on the proportion of the population hospitalized across ages (depicted as *P*_*H*_). To illustrate this, we first focus on a scenario in which *κ* = 0.5 as an example (Fig. 3; Fig. S5). We find that increasing levels of social distancing can have different effects on *P*_*H*_, depending on the targeted groups. For example, targeting social distancing at elderly under an ‘adults first’ vaccination strategy reduces *P*_*H*_ more effectively than under an ‘elderly first’ strategy (compare trends in purple lines in Fig. 3). Nevertheless, the elderly first strategy has overall lower *P*_*H*_. In addition, under ‘elderly first’, a uniform or adult social distancing is preferred. A comparison to simulations in which the probability of hospitalizations (*h*_*j*_) is uniform across all age groups indicates that age-dependent disease severity is a major factor underlying these patterns (Fig. S6).

**Fig 3.**
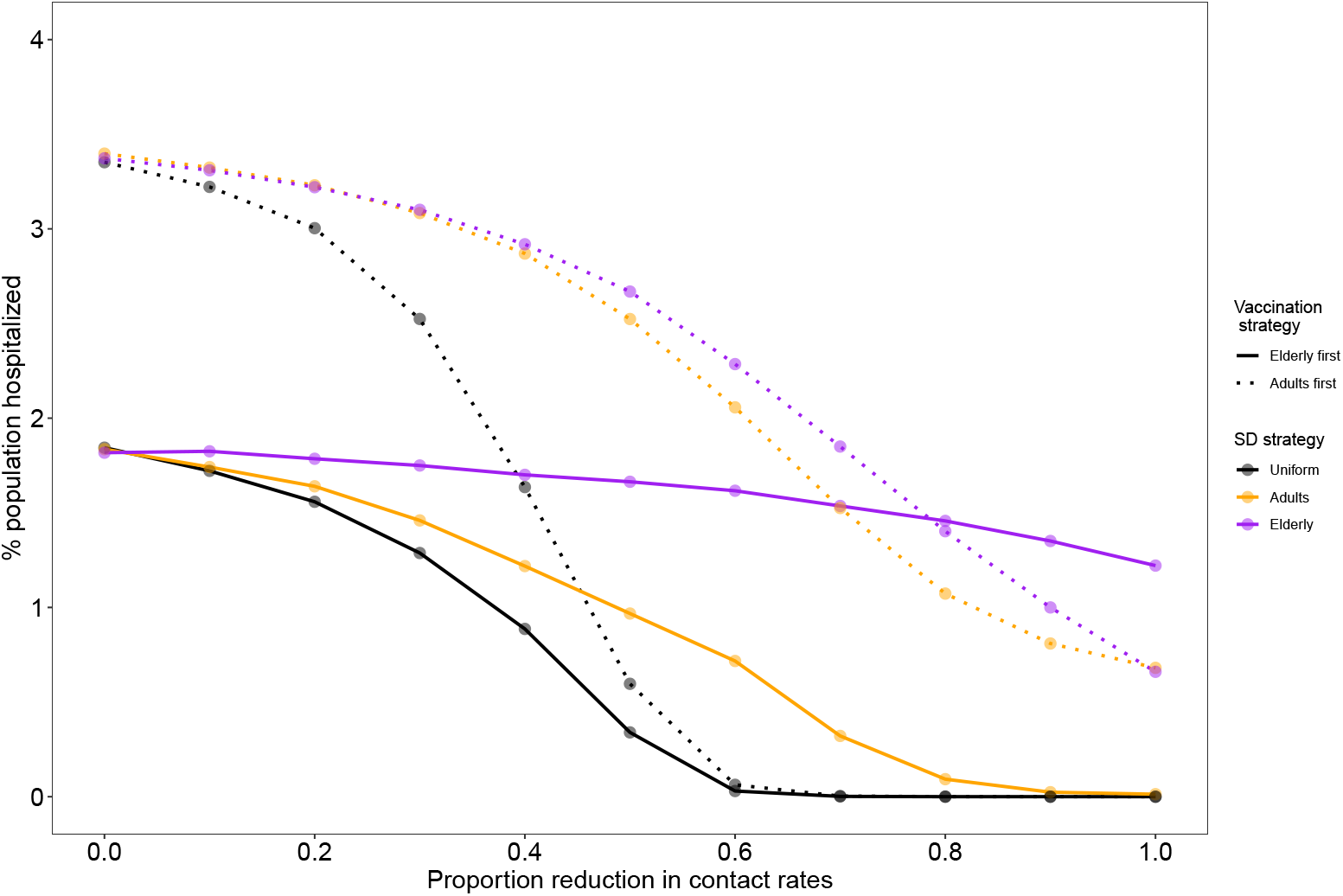
Effects of joint interventions on the proportion of the population hospitalized (*P*_*H*_). Each data point represents a combination of vaccination strategy (solid vs. dotted lines), social distancing strategy (colors) at particular social distancing strength (x-axis). Simulations were run for daily deployment of *κ* = 0.5% of the population.

To explore which vaccination strategy works better under different social distancing strategies in a systematic manner, we measured the difference in number of hospitalizations between the vaccination strategies as

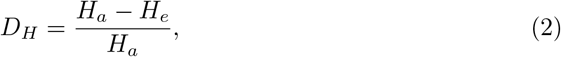

where *H*_*e*_ and *H*_*a*_ are the total (across ages) number of hospitalizations under the elderly first or adult first strategies, respectively. For example, if *H*_*e*_ = 15, 000 and *H*_*a*_ = 20, 000 then *D*_*H*_ = 25%. Hence, *D >* 0 indicates that vaccinating elderly is preferred to adults.

We detect a general trend in which an elderly first strategy is preferable when combined with a uniform or adult-targeted social distancing (Fig. 4). In tandem with our results on vaccine efficiency, under very strong social distancing targeted at the elderly, the effect of vaccination is overridden by that of social distancing, and vaccinating adults is preferred (Fig. 4B). Finally, an elderly first strategy is mainly preferred in high vaccination rates when social distancing is low (Fig. 4). This observation is crucial because it indicates that with low vaccination rates an elderly first strategy may have only a marginal benefit under some conditions. However, with moderate-high vaccination rates and social distancing targeted at adults, an elderly first strategy performs better than an ‘adult first’ strategy, even under low levels of social distancing.

**Fig 4.**
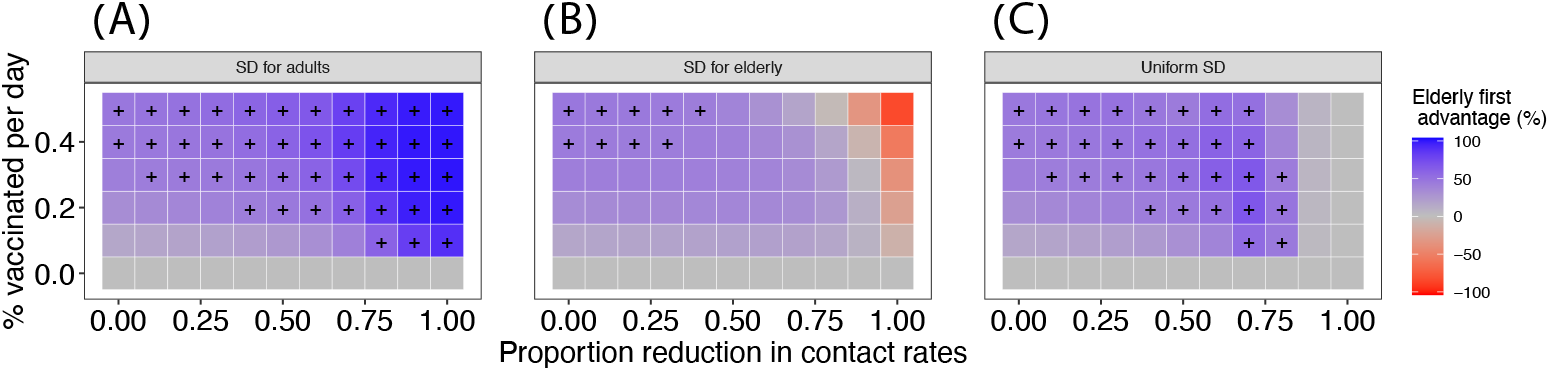
Systematic comparison between vaccination strategies in reducing hospitalizations. Each square in the heat map depicts the percent difference in the number of hospitalizations between the two vaccination strategies (*D*_*H*_ ; see text for equation) for a given strength of social distancing (x-axis), vaccine deployment rate (*κ*, y-axis) and social distancing strategy (panels A-C). Color scale depicts the advantage of an ‘elderly first’ strategy compared to an adults first strategy. Combinations higher than the median *D*_*H*_ are marked with a + sign.

### Reproductive number *R*_*t*_ depends on the combination of vaccination and social distancing

A key objective of interventions is to bring the number of active cases to a level that would prevent further transmission in the population. During the course of an epidemic, this is typically quantified using the reproductive number *R*_*t*_, which is the time-varying analog of *R*_0_. When *R*_*t*_ *<* 1 every person will infect on average less than one other person, indicating that the disease can no longer spread. We estimated *R*_*t*_ from simulation data using the method of [20] as recommended by [21]. This method is also used by the Israeli Ministry of Health. Briefly, *R*_*t*_ is calculated as

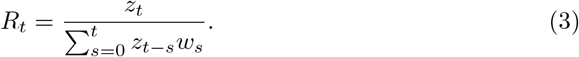

In empirical data *z*_*t*_ is the number of new infected individuals detected at day *t* but in our simulations we have full knowledge on active cases and use *z*_*t*_ = *I*_*t*_ + *A*_*t*_ + *P*_*t*_. The denominator is the sum of the infection potential of those that were infected within the previous *s* days. It is calculated using *w*_*s*_, which is a probability distribution for the interactivity profile, dependent on the time since infection. Following [22] and references therein, and the Israel Ministry of Health, we estimated *w*_*s*_ using a gamma distribution with a mean of 4.5 and standard deviation of 2.5, and calculated *R*_*t*_ over a period of *s* = 7 days.

Intuitively, the time it takes to reach *R*_*t*_ = 1 will be fastest when the disease is allowed to spread without any interventions because the amount of susceptible individuals decreases rapidly. We indeed observe this pattern (Fig. 5A). Nevertheless, this is an undesired situation that will cost many lives. The goal is therefore not necessarily to bring *R*_*t*_ below 1 as fast as possible, but to reduce it to prevent massive infections and resulting hospitalizations. Social distancing significantly increases the time it takes to reach *R*_*t*_ = 1 (Fig. 5) but lowers it. Vaccination positively interacts with social distancing, further reducing *R*_*t*_, but only when vaccination rates are high enough. In our model, a daily deployment to 0.1% of the population has only a marginal benefit, whereas that of 0.5% significantly reduces *R*_*t*_, especially when combined with a strong social distancing (Fig. 5C). Interestingly, under uniform social distancing, which is the most common strategy applied, *R*_*t*_ is little affected by the vaccination strategy (see Fig. S7 for other strategies).

**Fig 5.**
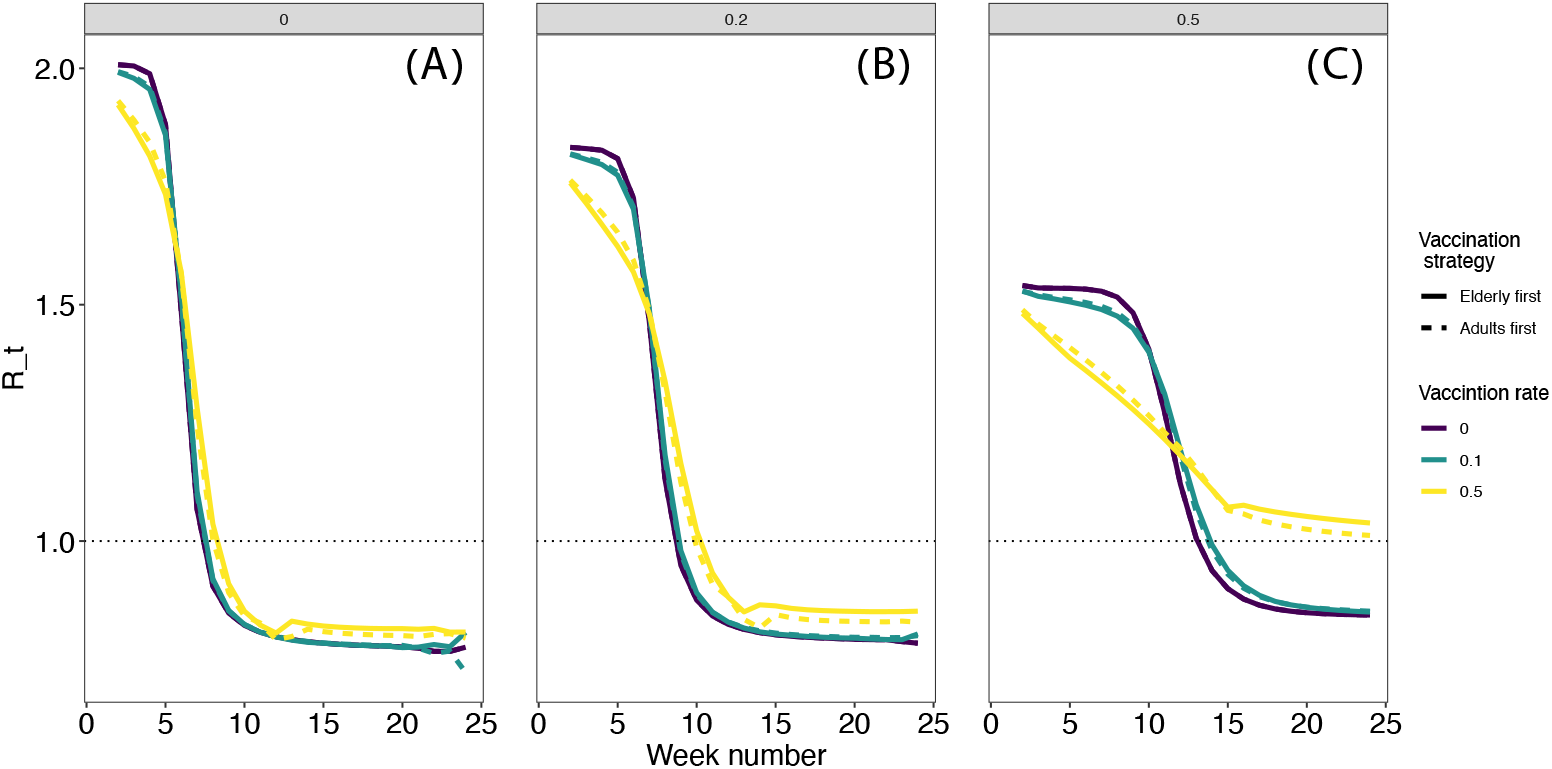
*R*_*t*_ for a uniform social distancing strategy. Each column represents a proportion of reduction in contact rates: 0 (no social distancing, panel A), 0.2 (B), and (C). *R*_*t*_ was calculated weekly (x-axis) as descried in the main text. Other social distancing strategies are presented in Fig. S7.

## Discussion

We used a deterministic compartmental model to mechanistically explore the qualitative interplay between vaccination and social-distancing on COVID19 disease dynamics. Strong social distancing keeps the number of susceptible individuals high but without a vaccine, infection and concomitant hospitalizations are inevitable. From a public health standpoint, social distancing and vaccination are short- and long-term interventions, respectively. Hence, a combination of these should provide greater benefits than when applied separately. We find that synergism between interventions can indeed be obtained via particular combinations. For example, by prioritizing vaccines to elderly and applying social distancing to adults. However, there are also quantitative effects. Most notably, under low vaccination rates the elderly first strategy that has been found to be preferable (e.g., [12, 14]) may have only a marginal benefit over an adult-first strategy, including for bringing *R*_*t*_ below 1 but increasing social distancing can increase this vaccination strategy’s efficiency.

Modeling is instrumental to informing policy makers and the public about possible scenarios of disease progression and the potential efficacy of different intervention methods [18, 23–25]. Estimation of the parameters necessary for accurate modeling is improving considerably, including social parameters. For example, a recent study showed only about 70% vaccine acceptance in Israel [19]. In our simulations we found no qualitative difference in effect on hospitalizations between 70%, 80% and 100% acceptance (see sensitivity analyses) because deployment to individuals with vaccine hesitancy is obtained when infections may be close to their peak. An effect of vaccine hesitancy may be notable under higher rates or vaccination or within other modeling frameworks. Researchers should recognize limitations of this kind and that inaccurate parameterization and ignoring model assumptions (e.g., geographical heterogeneity) could potentially lead to erroneous conclusions [26]. Our model assumes homogeneous mixing in space and reflects an average population—for example, it does not include household, school, or work dynamics, which are relevant when investigating epidemic infections via contacts. Therefore, results should be considered qualitatively to provide general guidelines.

Nevertheless, when interpreted within the known limitations, our study provides valuable insights into the joint effect of vaccination and social distancing and the mechanisms underling their interplay. It also provides initial guidelines for policy makers. For example, for some strategy combinations, vaccine efficiency varies little with increasing strength of social distancing, indicating that the social burden of social distancing may be relaxed. In addition, despite the synergism with social distancing, vaccination has little effect on *R*_*t*_ when deployed to small proportion of the population. This is because a vaccine is deployed during the pandemic, while non-vaccinated individuals continuously get infected. Here we considered deployment rates typical for the world average but the effect of vaccine should more notable with higher deployment rates. In contrast to vaccine that reduces the number of susceptible individuals, social distancing increases the time to reach *R*_*t*_ = 1 because it only impedes infections. Therefore, social distancing is a strategy to “flatten the curve” but, as we show here, the way that it is applied and combined with vaccinations is important.

Our model contains, beyond an age structure and age-dependent contact matrices, all the main states of COVID19 and its implementation allows for flexible prioritization strategies: One can choose a certain order of age groups to vaccinate, and if the number of individuals to vaccinate at a certain group is depleted but there are still vaccines available, the model shifts dynamically to the next group. While these features have been used in separate studies, they have not been combined in a single framework of deterministic modeling. Our model is therefore an excellent tool to mechanistically test a range of hypotheses and strategies for interventions. For example, we did not include children in our targeted interventions because vaccine is not available for them (yet) and to reduce the complexity of the study design. This is, however, a necessary future direction.

Vaccine prioritization is an ethical issue to which some guidelines have been laid out [8, 9]. We advocate for a mechanistic understanding of the factors that affect vaccination success and how to combine interventions in a way that will align with ethical guidelines. Our study provides insights towards these goals.

## Methods

### Ethics statement

All the data we use originates from publicly available resources, such as academic literature or online epidemiological sources. We do not use any clinical data that requires IRB.

### Model structure

The model is a mass action model that reflects a situation in which the number of contacts is independent of the population size–a reasonable choice for directly-transmitted diseases [27]. It follows a population of *N* individuals, divided into nine age groups, depicted with the subscript *j* (Table S1) and is described in the following equations:

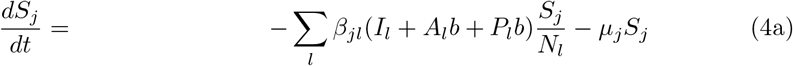

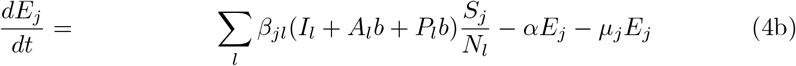

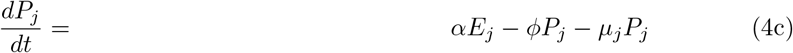

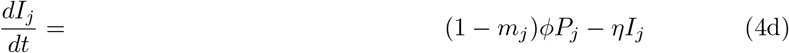

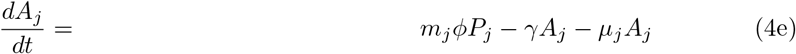

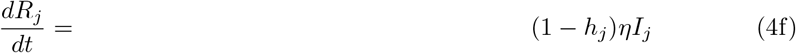

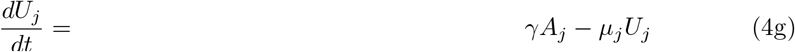

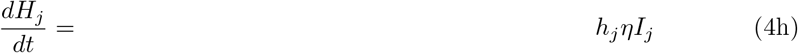

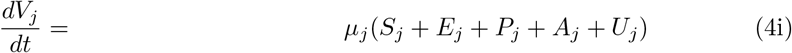

### Model states and parameters

Parameters for this kind of model are of two kinds. First, “biological” parameters, the variation of which is less related to country-specific demographics. These include, for example, recovery time (*γ*), the probability of developing an asymptomatic disease (*m*), incubation period (*α*), time spent in a presymptomatic stage (*ϕ*) and age-dependent probability of hospitalization (*h*). We obtained these parameters from the literature (see below). Second, demographic “social” parameters that may vary between countries and include age structure and contact rates between age groups. We obtained age structure from https://unctadstat.unctad.org/wds. Below we detail model states and the parameters we use to transition between them (see Table S2 for a summary).

#### Infected states

Individuals start at a susceptible state (*S*) and become infected upon an encounter with infectious individuals that are either presymptomatic (*P*), symptomatic (*I*) or asymptomatic (*A*). Following [17], we include a “scaling parameter”, *b*, ranging between 0-1 that determines the degree of infectiousness of states *A* and *P* compared to *I* (1 means a/pre-symptomatic are as infectious as symptomatic). We present results with *b* = 0.5 [17] but our results do not change qualitatively with other values. Infections occur at a rate specified by the infection matrix *β*_*lj*_, which we calculated as the product of the infection probability *q* and contact rates between age groups *j* and *l*, given by the contact matrix *C*_*lj*_. Therefore, *β*_*lj*_ = *q · C*_*lj*_ and we now detail how we obtained *C*_*lj*_ and *q*.

#### Infection parameters

We obtained *C*_*lj*_ for different countries from [28], which is currently the most comprehensive empirical (not inferred, e.g. [29]) survey of contacts relevant for the transmission of diseases such as COVID19, using the R package socialmixr. These contact matrices describe the daily mean number of contacts that participants have with people at different age groups (Fig. S8). We used the Italian contact network for Israel as a one for Israel is not available but results were qualitatively consistent across countries, including Italy. Mossong et al. [28] defined physical contacts as those that included interactions such as a kiss or a handshake and nonphysical contacts as those involving being in close proximity for a certain amount of time (e.g., a two-way conversation without skin-to-skin contact). Because SARS-COV-2 can also be transmitted via air droplets and aerosols, we included all physical contacts and those non-physical contacts that lasted for 15 minutes or more. Infection due to an encounter with individuals from the same age group occurs when *l* = *j*. We halved the matrix diagonal to avoid counting the same interaction twice.

#### Estimating infection probability, *q*

We calculated the average rate of infection 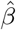 by fitting an exponential model of the form 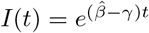 (sensu [30]) to Israeli case data across ages. This equation describes the invasion of the virus to a completely susceptible population at the early stages of the disease (days 1-35, before interventions were forced in Israel). This model gave a value of 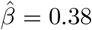 infections per day (*p <* 0.001, fit: *R*^2^ = 0.98). Estimating 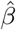 from *R*_0_ gave a similar estimate of 0.41 and this difference does not affect the results (see sensitivity analysis). Because 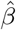 is an average across all the population, we calculated the infection probability given an encounter between a susceptible and infected individuals as 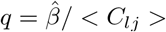. We then incorporated the recent estimates by [22] that children ages 0-20 are 43% less susceptible and 63% less infected than adults by reducing *q* for these ages proportionally.

#### Parameters of infection progression

Upon infection, the virus has an average latency period of 6.4 days (*α* = 1*/*6.4) [31] and individuals remain in presymptomatic stage for an average of 2.1 days (*ϕ* = 1*/*2.1) [32], after which individuals become either asymptomatic with a probability *m*_*j*_ or develop symptoms with probability 1 − *m*_*j*_. The fraction of asymptomatic COVID19 infections was estimated at 30-40% [33]. In the absence of data on age-specific asymptomatic infections [17], we have set *m* = *m*_*j*_ = 0.4 for all ages. However, our model retains the flexibility to quantify the role of age-dependent asymptomatic probabilities in COVID19 epidemiology (*m*_*j*_) because a previous study on corona viruses (not including SARS-COV-2) found that children tend to be less symptomatic than adults [34].

#### Parameters for infection outcomes

Asymptomatic individuals stop infecting (*U*) within 7 days (*γ* = 1*/*7) – an estimate from [35] that lies between that of [32] (mean 5 days) and that of [36] (mean 11 days). Results were qualitatively the same when we used the estimate by Davies et al [32], also used in [12]. We assume that symptomatic individuals are identified and removed to quarantine (*R*) within 1 day (*η* = 1). Depending on age, with probability *h*_*j*_ individuals can develop severe symptoms and be hospitalized (*H*). Therefore, *H*_*j*_ represents the cumulative number of hospitalized individuals at any given time. We calculated *h*_*j*_ using data from [37] as the fraction of hospitalized cases out of all cases (see Table S1 for details). This probability is not expected to vary greatly between countries as age-dependent hospitalization has a strong biological rather than social component in countries which are culturally similar. Effectively, individuals in states *V, U, R* and *H* cannot further infect. Separating between *R* and *U* is useful for scenarios in which quarantine should be considered separately from asymptomatic recovery (e.g., for economic reasons because some quarantined individuals cannot work).

### Interventions

#### Vaccination

We administer vaccines to people in states *S, E, P, A* and *U*, as they have not shown symptoms and are therefore considered susceptible from a public health perspective. We assume the vaccine prevents infection as well as disease, has an efficiency of 95%, and affects all individuals in the same way. Vaccines are deployed from the beginning of the simulation at a constant rate of *κ* vaccines per day, which we set as the percentage of the total population that a government can vaccinate a day. For example, in Israel (population about 8.7 million), vaccinating 0.2% of the population every day translates into deployment of about 17400 vaccines daily and over 1.5 million vaccines in the course of 90 days. We use values of *κ* ranging from 0.1% to 0.5%. The current rate at the EU, for example is 0.2-0.3%, and the world average is 0.1% (https://ourworldindata.org/covid-vaccinations).

In our simulations *κ* is first divided across the target age groups in proportion to the number of people left to vaccinate in each group (*κ*_*j*_). Then, the rate of vaccination (vaccines per person per day) is calculated as:

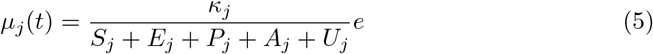

where *e* is vaccine efficiency, set to 95% [38].

#### Social distancing

Social distancing is the act of reducing contacts. Hence, we define 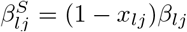, where elements of *x*_*lj*_ range between 0 (no social distancing) and 1 (no contacts whatsoever), and act to reduce contacts within and between groups. *x*_*lj*_ is the quantity depicted on the x-axis of figures (e.g., Fig. 2). We note that the multiplication *x*_*lj*_*β*_*lj*_ is an element-by-element multiplication rather than a matrix multiplication and that intervention is symmetric. For example, *x*_1,1_ = 0.4 will reduce contacts between juveniles within the age group 0-9 to 60% of the non-intervention level and *x*_3,1_ = *x*_1,3_ = 0.9 is a strong social distancing intervention reducing contacts between age groups 0-9 and 20-29 to 10% of their non-intervention level.

### Model implementation and sensitivity analyses

We implement the mathematical model with the package deSolve in R [39]. To facilitate future studies and study replication, we have developed a pipeline that allows changing parameters and running the model on High Performance Computing systems or locally with relative ease. This is described in the GitHub repository.

We run simulations for 24 weeks and have conducted multiple sensitivity analyses for the following parameters: *γ, α, ϕ, η, b*, and the proportion vaccinated (70% and 100% acceptance). For *β*, we tried an alternative estimation by calculating the *R*_0_ for Israel (across ages) using the method of [40] implemented in the R package R0 [41]. We used case data on days 1-35, before interventions were forced because *R*_0_ describes the invasion of the virus to a completely susceptible population at the early stages of the disease. This analysis gave an *R*_0_ = 2.9. We then calculated 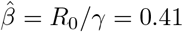 [27]. Qualitatively, our results for all sensitivity analyses were robust to changes in these parameters. We also repeated our analyses for Belgium, Germany and Italy using *R*_0_ estimates from [42], and obtained similar results. Finally, calculating *R*_*t*_ using the method of [40] did not change the results qualitatively. Outputs of all these simulations can be found in the GitHub repository, along with the code.

## Data Availability

This is a simulation study. Code will be published upon acceptance in the peer-reviewed journal

## Code Availability

All analyses were performed in R (version 4.0) and the code is fully available in https://github.com/Ecological-Complexity-Lab/COVID19_vaccine_model.

## Acknowledgments

We thank Profs. Yoav Tsori and Roni Granek for comments on a previous version of the model. This research was supported by the Ministry of Science & Technology, Israel (grant no. 3-16893).

---

**Table S1.** Population structure of Israel and relevant parameters. *j* is the ID of the age group; Proportion is the proportion of the age group in the Israeli population; *h*_*j*_ is the probability of hospitalization taken from [1]. We preferred this data source because of its high resolution and high number of cases that provides better accuracy for calculating proportions. This probability is not expected vary greatly between countries as age-dependent hospitalization has a strong biological rather than social component in countries which are culturally similar. Indeed, we contrasted it with Israeli data from [2] and proportions were highly similar.

| $j$ | Age | Proportion | $h_j$ |
| --- | --- | --- | --- |
| 1 | 0-9 | 0.20 | 0.024 |
| 2 | 10-19 | 0.16 | 0.014 |
| 3 | 20-29 | 0.14 | 0.021 |
| 4 | 30-39 | 0.13 | 0.038 |
| 5 | 40-49 | 0.12 | 0.058 |
| 6 | 50-59 | 0.09 | 0.091 |
| 7 | 60-69 | 0.08 | 0.166 |
| 8 | 70-79 | 0.05 | 0.296 |
| 9 | 80+ | 0.03 | 0.451 |

**Table S1.** Study parameters. The proportion of the target population accepting vaccination was applied computationally and does not appear in the model equations. The values noted are those used in the main simulation (experiment 1). We also conducted sensitivity analyses. The complete set of experiments that includes all values for all parameters can be found in the GitHub repository (file experiments.csv).

| Parameter | Type | Value | Description | Reference |
| --- | --- | --- | --- | --- |
| $\beta$ | rate | 0.31 | infections per person per day | |
| $\alpha$ | rate | 1/6.4 days | incubation time | [1] |
| $\gamma$ | rate | 1/7 days | recovery rate | [2] |
| $\phi$ | rate | 1/2.1 days | time in presymptomatic stage | [3] |
| $\eta$ | rate | 1 day | time to quarantine upon showing symptoms | |
| e | proportion | 0.95 | vaccine efficiency | [4] |
| b | proportion | 0.5 | relative infectiousness of presymptomatic and asymptomatic relative to symptomatic | [5] |
| h | proportion | Table S1 | Proportion of cases hospitalized | [6, 7] |
| m | proportion | 0.4 | proportion of asymptomatic infections | [8] |
| p | proportion | 0.8 | proportion of target population accepting vaccines | [9] |

**Figure S1:**
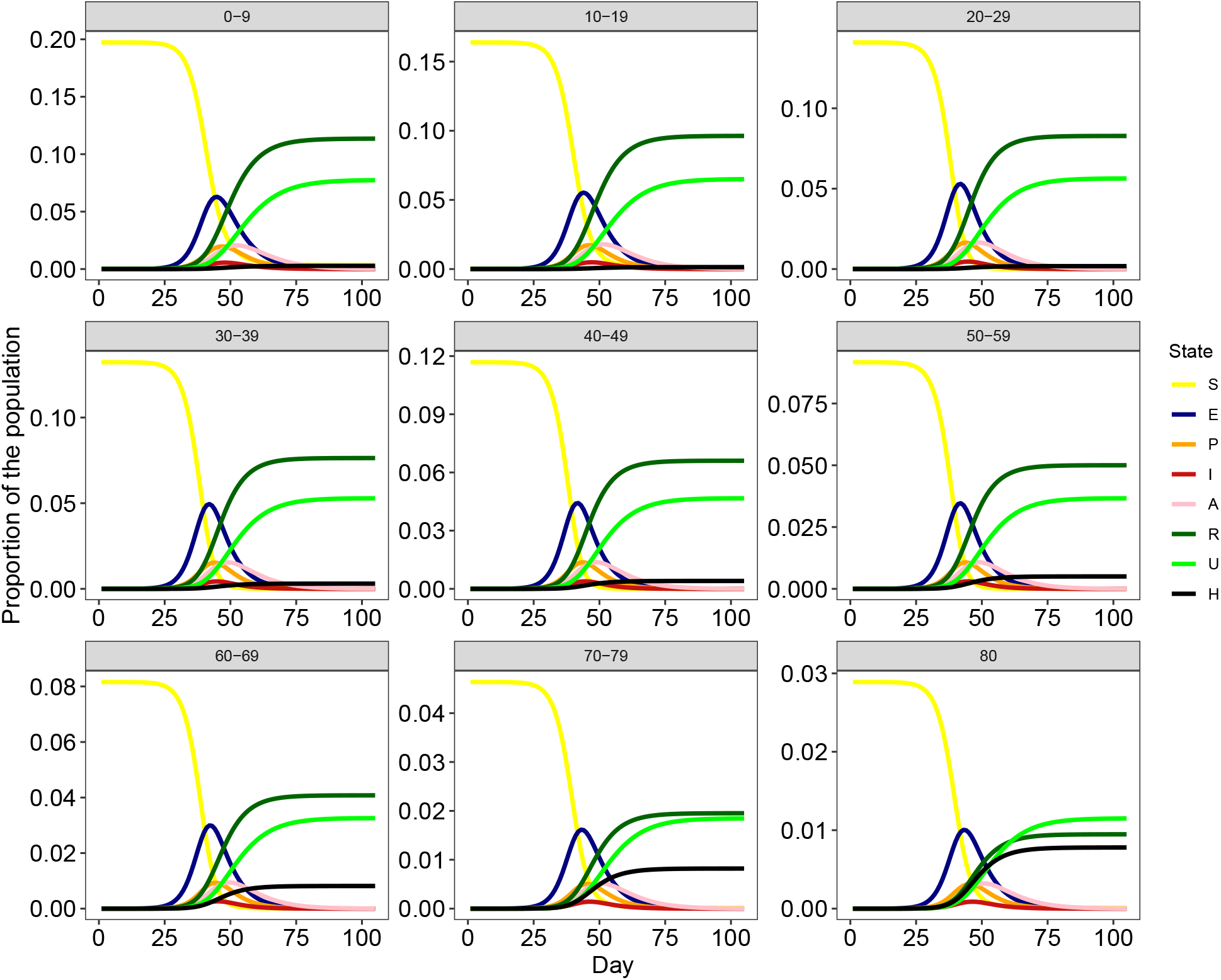
Epidemiological curves without vaccine. Epidemiological curves for the baseline model, with no vaccination or social distancing. The model follows a population divided to nine age groups as depicted in the figure. *S*: susceptible; *E*: exposed; *P* : presymptomatic; *I*: infectious and symptomatic; *A*: infectious and asymptomatic; *R*: removed to quarantine and then recovered naturally, and immune; *U* : recovered naturally, and immune; *H*: hospitalized.

**Figure S2:**
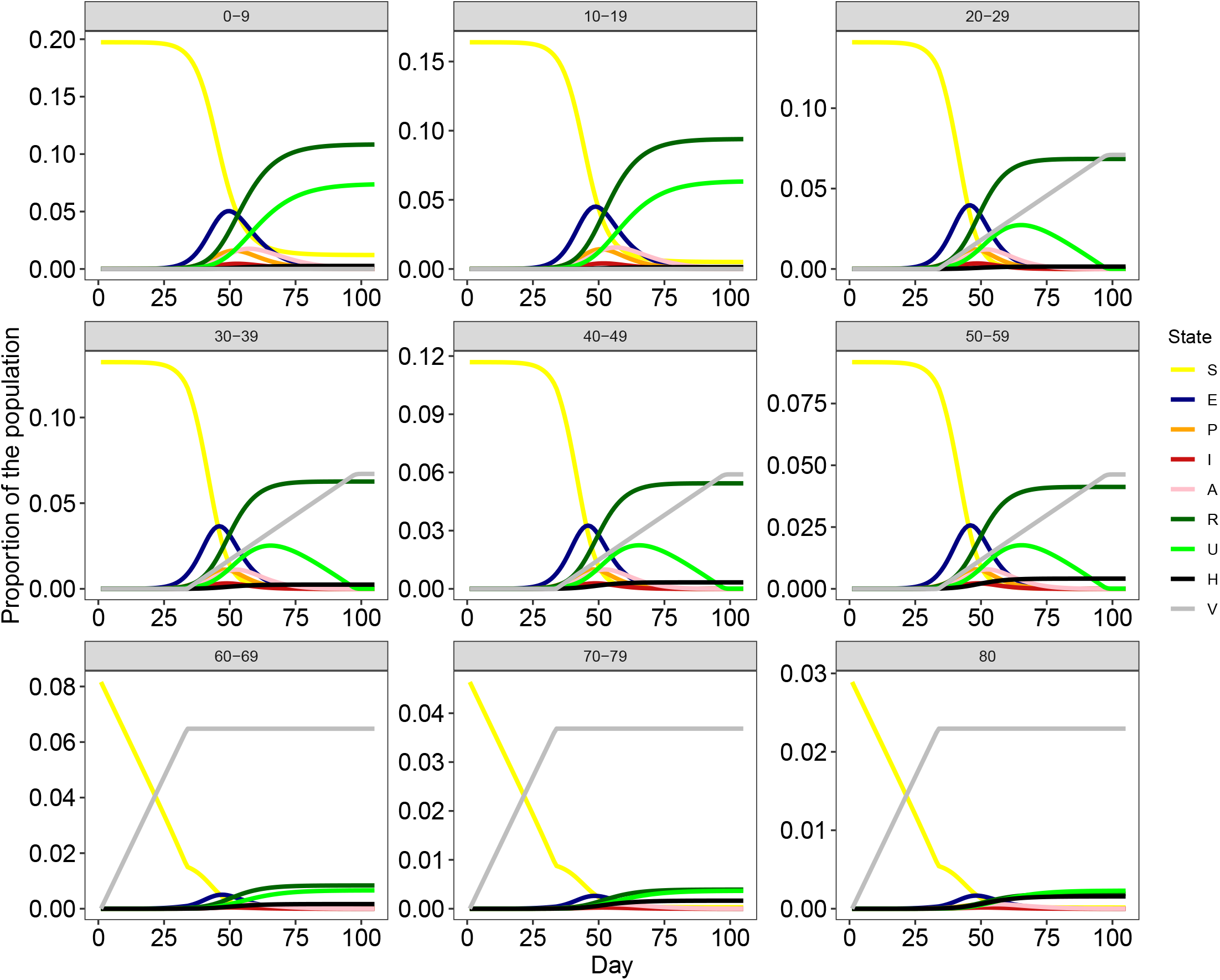
Epidemiological curves with vaccine. Epidemiological curves with vaccination (*κ* = 0.2) and no social distancing. The model follows a population divided to nine age groups as depicted in the figure. *S*: susceptible; *E*: exposed; *P* : presymptomatic; *I*: infectious and symptomatic; *A*: infectious and asymptomatic; *R*: removed to quarantine and then recovered naturally, and immune; *U* : recovered naturally, and immune; *H*: hospitalized; *V* : vaccinated.

**Figure S3:**
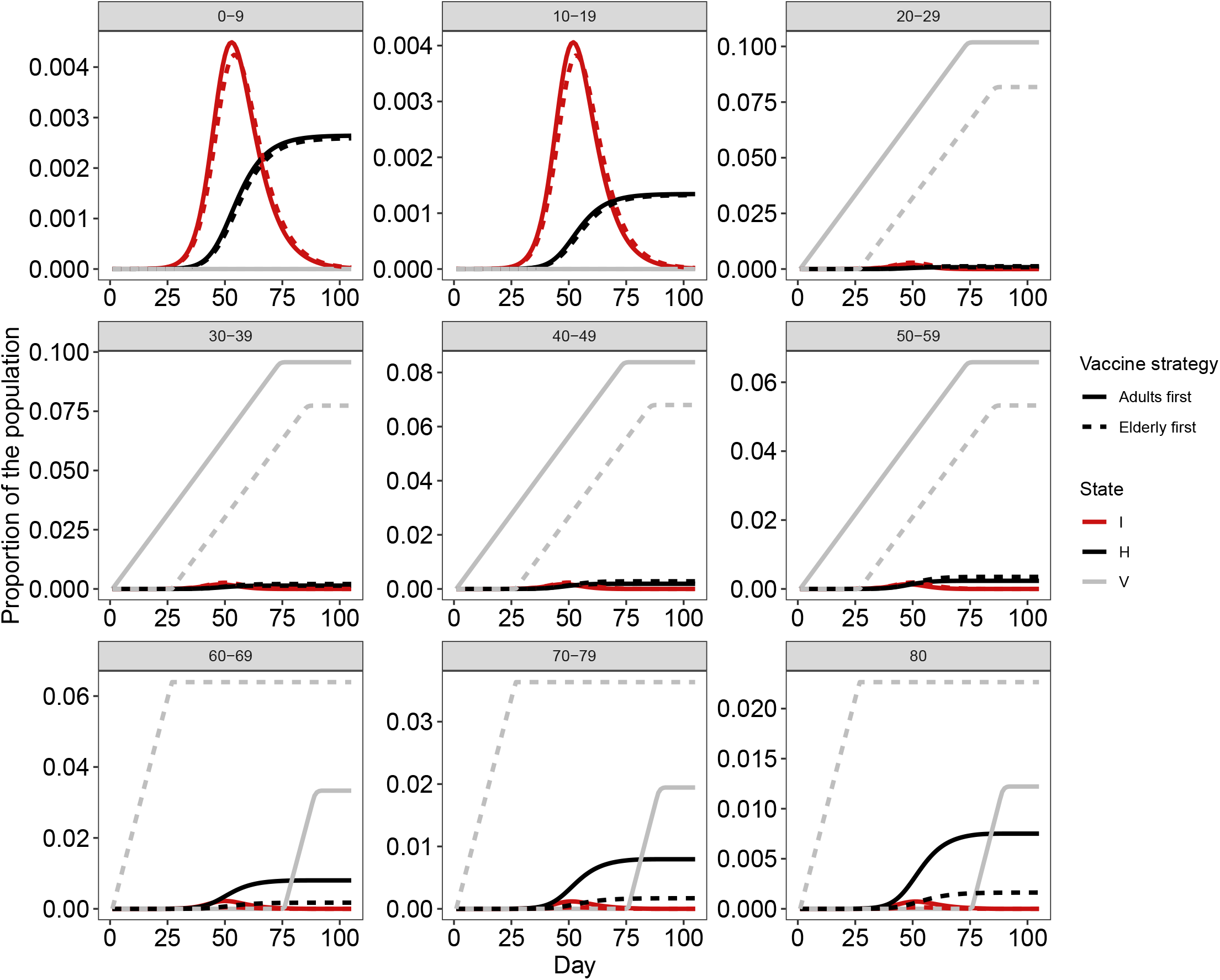
Comparison of dynamics between two prioritization strategies. Dashed and solid lines depict two strategies, respectively: (i) vaccinating all elderly (60+) and then all adults (20-59) and (ii) vaccinating all adults and then all elderly. This example is for *κ* = 0.5 (0.5% of the population is vaccinated per day) with no social distancing. In the first strategy, around 35 days there are no more elderly to vaccinate and the model shifts to vaccinating adults (gray dashed line). In the second strategy, the switch to vaccinating elderly occurs after about 60 days because the number of adults in the population is larger than that of elderly. Most of the reduction in hospitalizations is obtained when the elderly are prioritized (compare black lines). *I* : infectious and symptomatic (red); *H* : hospitalized (black); *V* : vaccinated (gray).

**Figure S4:**
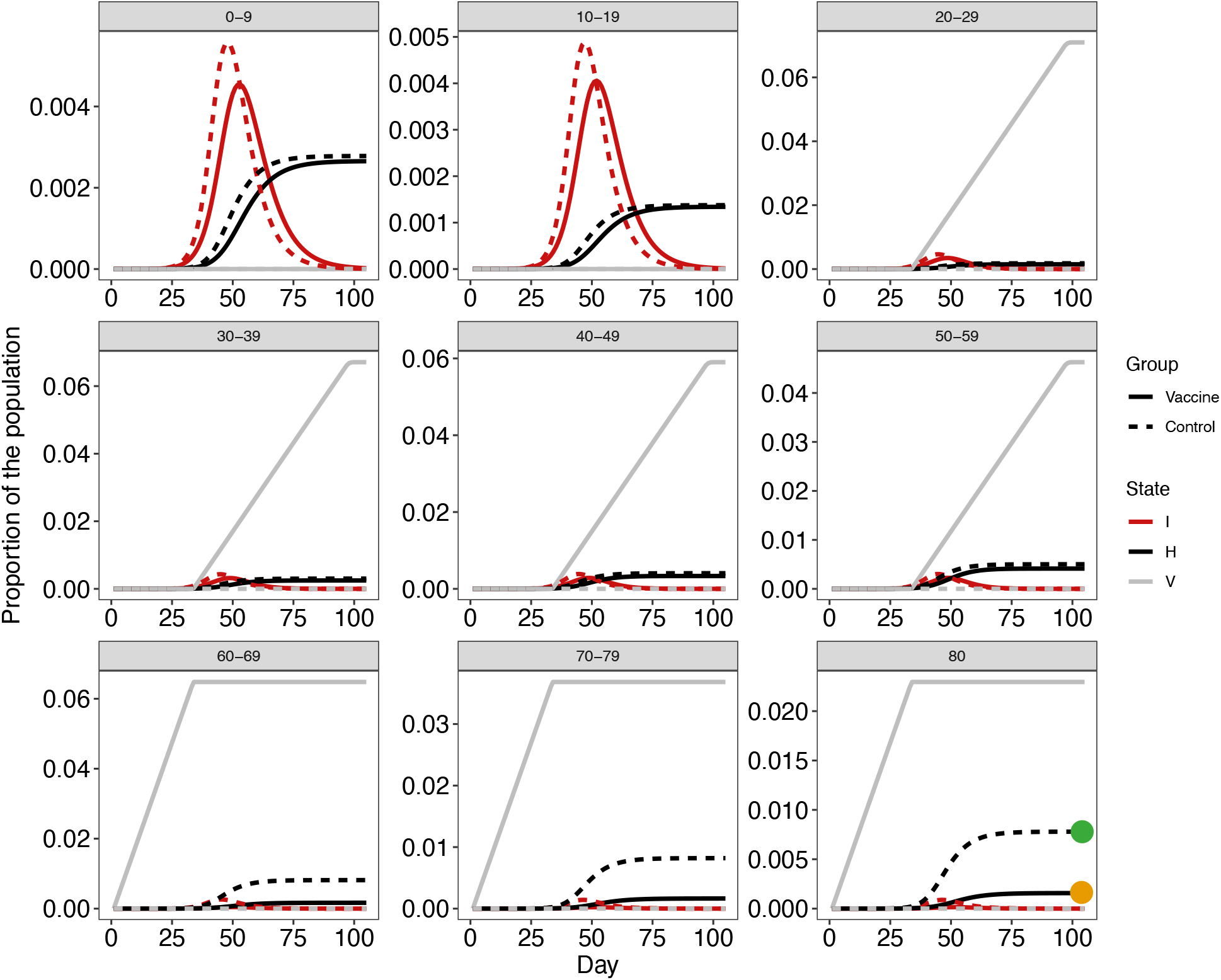
Comparison of control to vaccine intervention. Dashed lines depict control (no vaccine). Solid lines depict a strategy in which we first vaccinate elderly (60+) and then adults (20-59). In this example we use a single value of *κ* = 0.4 (0.4% of the population is vaccinated per day) and there is no social distancing. Around 20 days there are no more elderly to vaccinate and the model shifts to vaccinating adults (gray solid line). It is clear that most of the reduction in hospitalizations is obtained for the elderly groups (compare dashed to solid black lines). The orange and green dots mark the *H*_*j*(*κ*=0.2)_ and 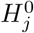 values (for *j* = 80+) used in equation 1 in the main text, respectively. Model was run for 30 weeks but 12 weeks are plotted here for clarity. *I* : infectious and symptomatic (red); *H* : hospitalized (black); *V* : vaccinated (gray).

**Figure S5:**
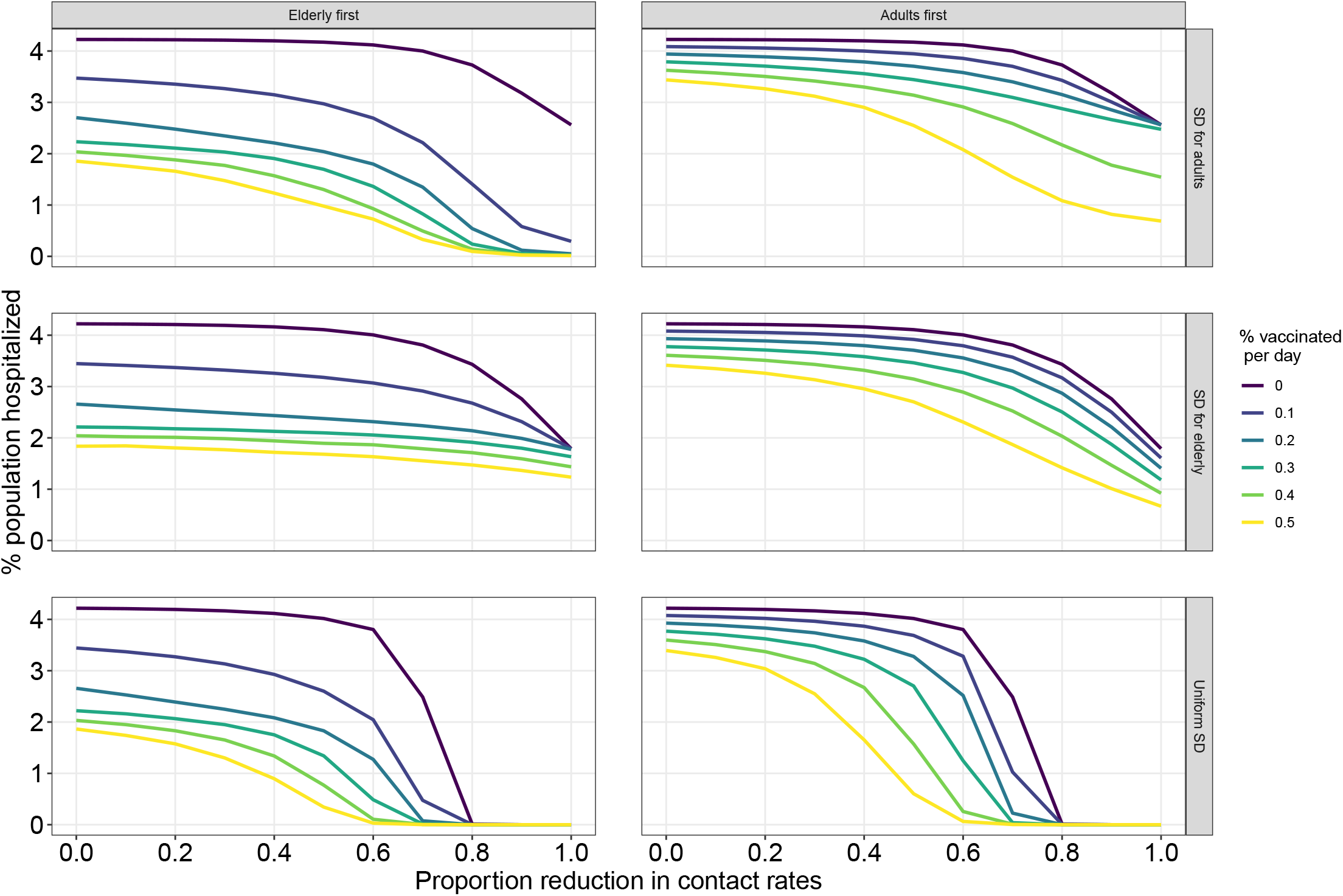
Effect of joint interventions on vaccination efficiency on proportion of hospitalization. The plot depicts *P*_*H*_ (y-axis) as a function of strength of social distancing (x-axis), vaccination rates (*κ*; line colors), vaccination strategies (columns) and social distancing strategies (rows).

**Figure S6:**
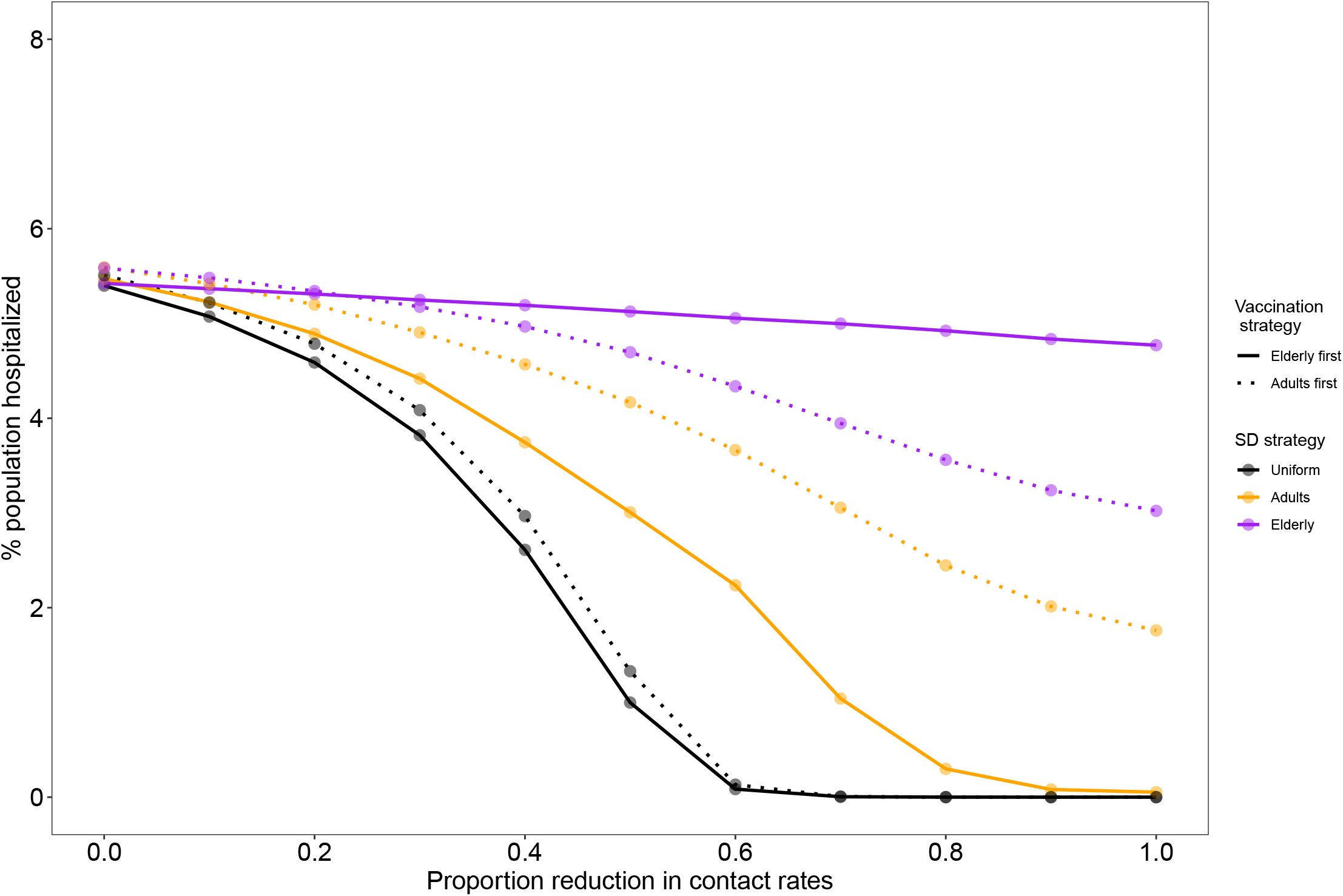
Effects of joint interventions on the proportion of the population hospitalized (*P*_*H*_) using age-uniform hospitalization probabilities. Each data point represents a combination of vaccination strategy (solid vs. dotted lines), social distancing strategy (colors) at particular social distancing strength (x-axis). Simulations were run for daily deployment of *κ* = 0.5% of the population. The probability of hospitalization was equal to the mean probability across ages (i.e., mean of *h*_*j*_)

**Figure S7:**
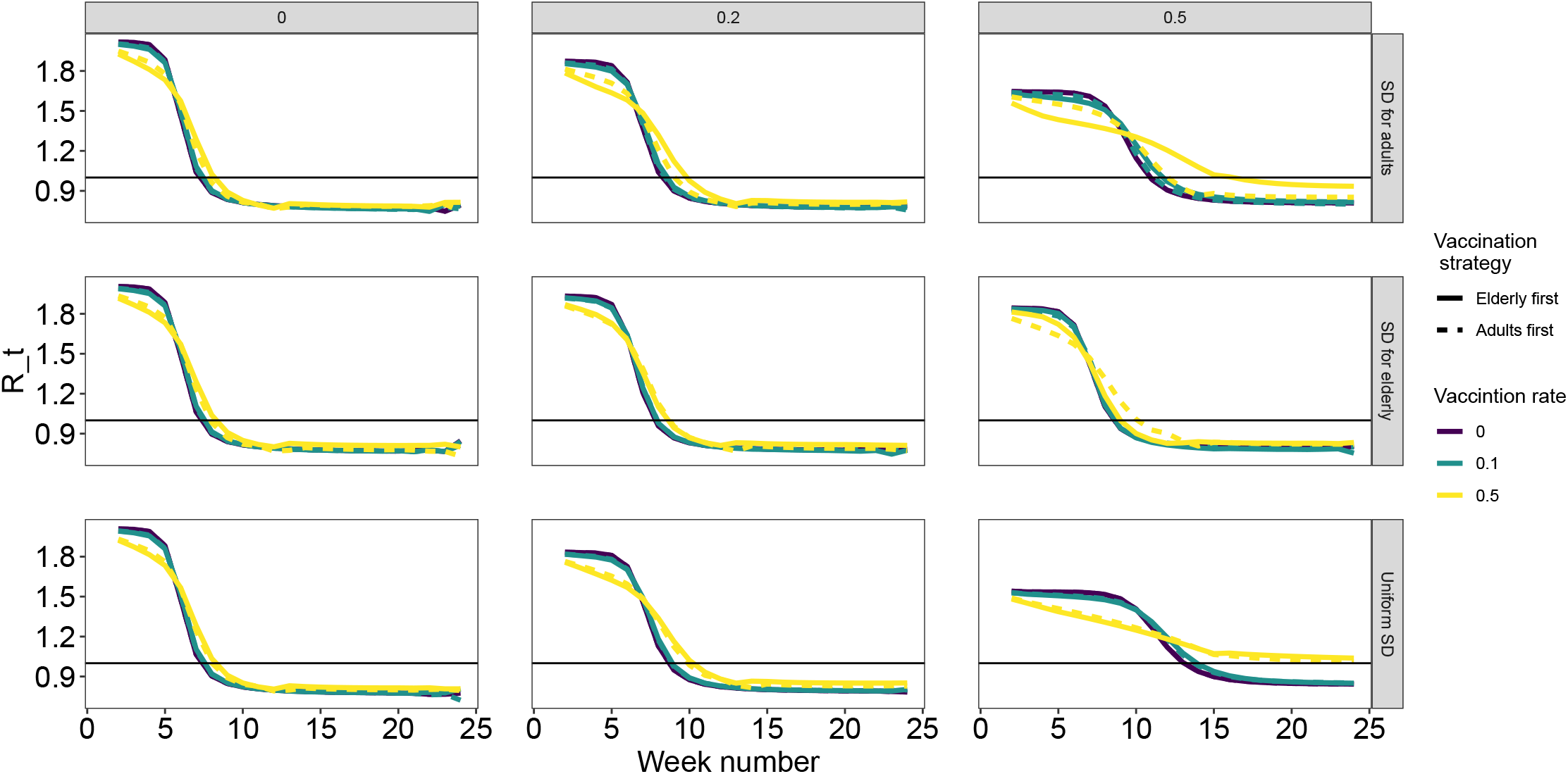
*R*_*t*_ for different social distancing strategies. Columns represent three levels of proportion of reduction in contact rates: 0 (no reduction), 0.2, and 0.5. Each row is a social distancing strategy. *R*_*t*_ was calculated for 7-day periods as descried in the main text.

**Figure S8:**
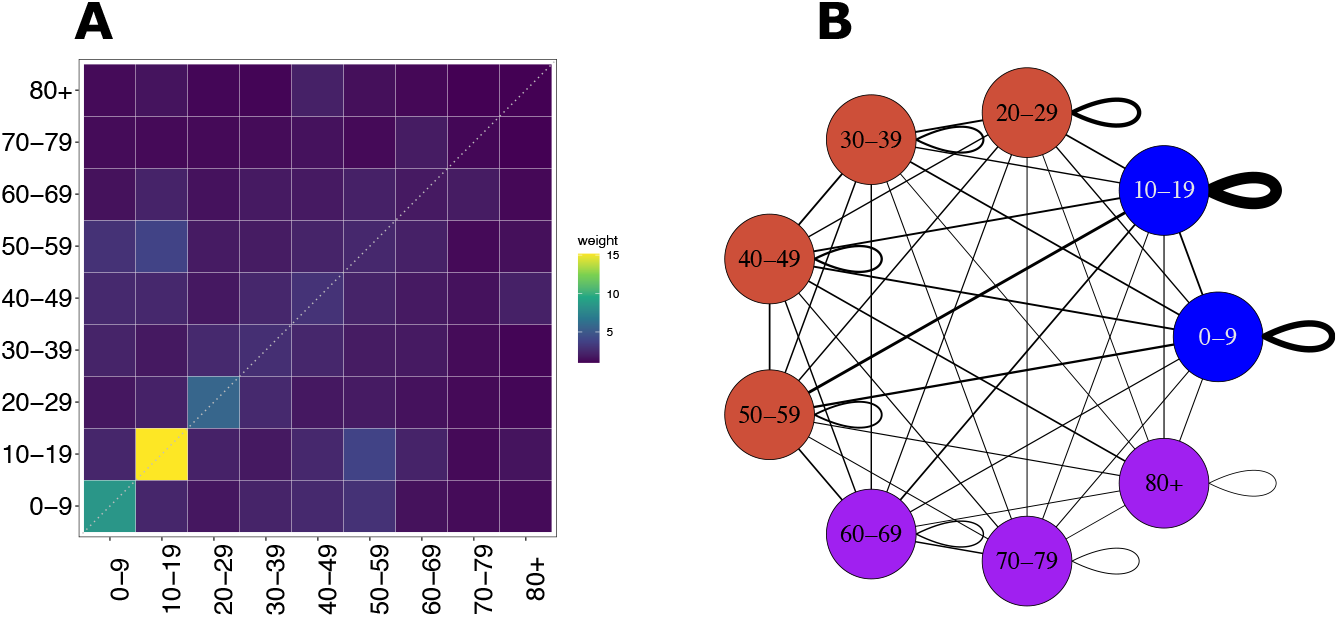
An example of an age-dependent contact rates. (A) Contact matrix for Italy calculated from data collected by Mossong et al. (2008). Matrix cells depict the mean number of daily contacts between people in different age groups. Diagonal cells depict contacts between individuals from the same age group. (B) The same data as in (A), represented as a network of contacts. Edge widths depict contact rates.

